# Bile acid dysmetabolism in Bangladeshi infants is associated with poor linear growth, enteric inflammation, and small intestine bacterial overgrowth

**DOI:** 10.1101/2025.02.04.25321650

**Authors:** Farah Hasan, Phillip B Hylemon, Rashidul Haque, William A. Petri, Abu S.G. Faruque, Beth D. Kirkpatrick, Masud Alam, Tahsin Ferdous, Talat Shama, Brett Moreau, Girija Ramakrishnan, Huiping Zhou, Alden Chesney, Felix Medrano Garcia, Ekaterina Smirnova, Preethi Prem, Yingsi Huang, Rashmi Bojja, Anubhav Thapaliya, Jeffrey R. Donowitz

**Affiliations:** Virginia Commonwealth University School of Medicine, Richmond, VA USA; Department of Microbiology and Immunology and Stravitz-Sanyal Institute for Liver Disease & Metabolic Health, Virginia Commonwealth University, Richmond, VA USA; Infectious Diseases Division, the International Centre for Diarrhoeal Disease Research, Bangladesh (icddr,b), Dhaka, Bangladesh; Division of Infectious Diseases & International Health, University of Virginia, Charlottesville, VA USA; Centre for Nutrition & Food Security, International Centre for Diarrhoeal Disease Research, Bangladesh (icddr,b), Dhaka, Bangladesh; Department of Microbiology and Molecular Genetics, The University of Vermont Larner College of Medicine, Vaccine Testing Center, Burlington, Vermont, USA; Division of Clinical Pathology, Virginia Commonwealth University Medical Center, Richmond, Virginia, USA; Richmond Veterans Medical Center, Richmond, VA, USA; Department of Biostatistics, School of Medicine, Virginia Commonwealth University, Richmond, VA USA; Department of Biostatistics, School of Medicine, Virginia Commonwealth University, Richmond, VA USA. Carle Illinois College of Medicine, Champaign, IL, USA; Virginia Commonwealth University, Richmond, VA USA. Carle Illinois College of Medicine, Champaign, IL, USA; Department of Management Science Miami Herbert Business School, University of Miami Coral Gables, FL; Edward Via College of Osteopathic Medicine – Virginia Campus, Blacksburg, VA USA; University of Texas Southwestern Medical School, 5323 Harry Hines Blvd, Dallas, TX, 75390, USA; Division of Pediatric Infectious Diseases, University of Virginia, Charlottesville, VA USA

## Abstract

**Introduction:** Environmental enteric dysfunction (EED) is a subclinical condition caused by fecal-oral contamination leading to enteric inflammation and dysbiosis. This study investigates bile acid metabolism in Bangladeshi children with EED and its association with growth impairment.

**Methods:** We conducted a cross sectional study of 100 Bangladeshi infants (aged 6-9 months) and quantified serum and fecal bile acids using LC-MS/MS. We compared profiles to a control group of 6 American children (6 – 12 months) and 80 older Bangladeshi children (aged 2 years).

**Results:** Bangladeshi infants had higher levels of plasma unconjugated primary (65.23% vs. 44.25%, p = .003) and sulfated primary bile acids (12.98% vs. <0.001%, p = .01), with lower primary conjugated bile acids (0.69% vs 2.74%, p = <0.001) compared to American children. Stool unconjugated primary bile acids were inversely associated with weight-for-age (regression coefficient [β] = −0.01, p = 0.01) and height-for-age z scores (β = −0.01, p = 0.03). Conjugated secondary bile acids were inversely associated with small intestine bacterial overgrowth (β = −1096.68, p = 0.05). Fecal myeloperoxidase was associated with sulfated secondary bile acids (β = −0.40, p = 0.04). Compared to 2-year-old children, the Bangladeshi infant’s serum had higher levels of unconjugated primary bile acids (65.23% vs. 9.20%, p = <0.001) and lower levels of primary conjugated bile acids (0.69% vs 80.38%, p = <0.001).

**Discussion:** Our data suggests an age-dependent defect in conjugation of primary bile acids in Bangladeshi children with compensatory hydrophilic shunting. Additionally, bile acid profiles are associated with intestinal overgrowth.

## Background

Twenty-one percent of deaths in children under the age of five years can be attributed to malnutrition (11). Pediatric malnutrition in low and middle-income countries (LMICs) is multifactorial with contribution from food insecurity, inadequate access to clean water, and unsanitary living conditions resulting in chronic enteric pathogen exposure. Chronic carriage of enteric pathogens leads to environmental enteric dysfunction (EED), a sub clinical condition characterized by chronic gut inflammation, small intestinal injury, and small intestinal dysbiosis (9). EED has been associated with deficient nutrient absorption and systemic inflammation which led to linear growth shortfalls and poor neurodevelopmental outcomes (2). Constant exposure to fecal-oral contamination leads to enteric inflammation and subsequent dysbiosis which, in turn, leads to reduced barrier function, villous blunting, and a decreased crypt-to-villus ratio (22). These histological changes recovered when subjects were removed from areas of poor sanitation and hygiene (22). While intestinal biopsies are the gold standard for diagnosing EED, endoscopy is invasive and therefore various biomarkers of intestinal and systemic inflammation have been used as proxy measures of EED burden (6). Childhood EED has been associated with growth stunting, decreased weight, and poor neurodevelopmental outcomes (22).

Among children with EED, small intestine bacterial overgrowth (SIBO) is common. SIBO was present in 11% of 18 weeks old Bangladeshi infants and the prevalence increased to 30 – 40% in 1 to 2-year-olds (8). In this population SIBO was associated with markers of intestinal inflammation (7). SIBO in the low-income country setting is also associated with growth stunting and language delay, independently of inflammation (4,8,23). Despite the association of SIBO and poor growth being noted in several studies across several continents, the nature of this association and the pathogenesis of SIBO remains unclear. One study did, however, note that the SIBO microbiome led to decreased lipid absorption when transplanted from patients into an in vitro murine intestinal m-IcCl2 cell line (23).

EED associated gut dysbiosis has also been linked to abnormal bile acid patterns (19). Bile acids act as detergents in the intestine to facilitate lipid digestion and absorption. They also serve as signaling molecules that regulate numerous metabolic pathways associated with childhood growth (14). Furthermore, bile acids have been shown to play a role in host susceptibility to inflammation and serve as sensitive markers to hepatic and cholestatic function (26). Bile acid metabolism defects can lead to fat-soluble vitamin deficiency, malnutrition, neurologic impairments, and stunted growth (10). Lack of bile acid conjugation has even been linked to the action of the bacteria proliferating in the upper small intestine (17). There is a paucity of literature investigating the role of bile acid metabolism in low- and middle-income country children with EED and malnutrition although preliminary evidence suggests altered bile acid metabolism in this population (13,18–19,25–26). A study looking at children in rural Malawi reported that children with EED displayed altered bile acid metabolism from a young age, with a higher proportion of bile acids conjugated with taurine instead of glycine. Total serum bile acids were also approximately 12% lower in children with EED compared with children without EED which may be reflective of impaired reuptake due to injury in the ileum (19). The same study also inversely correlated levels of primary unconjugated bile acids with age in children with EED (19). Another study in undernourished Pakistani children with EED found elevated levels of serum bile acids when compared to well-nourished local American children (26).

Based on these previous studies, we hypothesized that stunted children with EED and SIBO have altered bile acid metabolism. To test this hypothesis, we conducted a cross-sectional analysis of Bangladeshi children and compared them to American controls, investigating bile acid patterns in both the serum and stool.

## Methods

We conducted a cross-sectional analysis of Bangladeshi toddlers investigating the association between SIBO, EED biomarkers, and bile acid profiles in both serum and stool. We enrolled 100 toddlers from the urban neighborhood of Mirpur in Dhaka, Bangladesh. Enrollment was from October 2017 through July 2018. The majority of the homes in Mirpur are mud brick construction. Crowding is common with a mean of 5 people living in 1.5 rooms per dwelling.

Uncovered sewers flow throughout the neighborhood with municipal water lines often running through these sewer channels. Due to the location of our study clinic, subjects tended to come from the lowest socioeconomic strata of Mirpur. Enrolled children were 6-9 months old with no known chronic medical problems other than mild wasting as defined by a weight-for-age Z (WAZ) score between −1 and −3 standard deviations (SD).

Blood was collected by a pediatric phlebotomist in our study clinic. The stool was collected in our study clinic by field assistants (FA). Maintaining proper cold chain, samples were transported to our laboratory within 4 hours where they were aliquoted and placed in −80°C. One aliquot was removed for batched biomarker analysis at the completion of the study. A separate aliquot was later shipped to the United States on dry ice where samples were again stored in −80°C until they were removed for analysis. The cold chain was monitored throughout the shipping process. We also conducted both a 2-hour dual sugar urinary lactulose-mannitol test by LC-MSMS and glucose-hydrogen breath test on subjects using methods on which we have previously published (7,15). Pediatric urine collection bags were attached to the patients and were allowed to return to regular diet 30 minutes after LM test solution ingestion. 2 ml of urine was collected at the 2-and-5-hour marks. The samples were then analyzed by the HPLC-MSMS system (15). Glucose-hydrogen breath testing was done by QuinTron BreathTracker SC gas chromatography which collected breath via an age-appropriate anesthesia mask attached at 20-min intervals for 3 hours. Patients fasted for 3 hours prior to and throughout testing but were allowed water. Children younger than 12 months fasted for 2 hours (7). SIBO AUC (i.e. area under the breath hydrogen curve) was calculated by summing the trapezoidal area under the glucose-hydrogen curve using methods previously published (8). These two tests were conducted within one week of each other but not on the same day. Stool and serum were collected at the same time as glucose-hydrogen breath testing. Biomarkers were tested by commercially available ELISA and included stool regenerating family member 1 beta (REG1B) [TechLab, Inc. Blacksburg, Va USA], stool myeloperoxidase (MPO) [ALPCO. Salem, NH USA], serum C-reactive protein (CRP) [ALPCO. Salem, NH USA], and serum soluble CD14 (SCD14) [R&D Systems, Minneapolis, MN USA]. Anthropometry was measured using calibrated infant scales and an infant measuring board by staff trained in the procedure on the day of breath testing. Z scores for weight-for-age (WAZ), height-for-age (HAZ), weight-for-height (WHZ), and BMI-for-age (BAZ) were calculated using World Health Organization software (WHOAnthro).

We also obtained discarded serum samples collected and stored in EDTA from the clinical laboratories at Virginia Commonwealth University Hospital in Richmond, Virginia USA. Only samples from 6-12-month-olds were screened. Samples were collected for clinical purposes and sent within minutes of collection to the clinical laboratory where they were processed within 1 hour and stored at 4°C for 3 days. Samples stored longer than 3 days are generally discarded. Once a sample was identified as marked for discard, having sufficient volume for bile acid testing, and from the appropriate age range, the electronic medical record was reviewed by our team (JD) to ensure the children had no known gastrointestinal or metabolic diagnoses associated with altered bile acid metabolism.

Serum from both Bangladeshi and American children, as well as stool from Bangladeshi children underwent bile acid and 7α-hydroxy-4-cholesten-3-one (C4) profiling. C4 is a bile acid precursor, serving as an indicator of overall bile acid production. The serum samples were processed and the composition and levels of individual bile acid metabolism were measured using a Shimadzu liquid chromatography/tandem mass spectrometric (LC-MS/MS) 8600 system as described previously (24).

We first analyzed the composition of bile acids in both the serum and stool samples from both cohorts. Individual bile acid levels were expressed as percentages of total bile acids detected, excluding C4 which was analyzed as a continuous variable. If a bile acid was undetectable, it was assigned a value of 0. If there were no detectable levels, bile acids were valued at a 0. Bile acids were grouped based on the bile acid metabolism pathway as follows: CDCA and CA were categorized as unconjugated primary bile acids, TCA, GCA, TCDCA, and GCDCA were categorized as conjugated primary bile acids, CDCA-3-S and CA-3-S were categorized as sulfated primary bile acids, UDCA, DCA, and LCA were categorized as unconjugated secondary bile acids, TUDCA, GUDCA, MDCA, GDCA, TLCA, and GLCA were categorized as conjugated secondary bile acids, 7keto-DCA, 7keto-LCA, isoDCA, isoLCA, allo-isoLCA, and 3keto-LCA were categorized as secondary bile acid metabolites, LCA-3-S and UDCA-3-S were categorized as sulfated secondary bile acids, and TwMCA, TaMCA, TbMCA, GbMCA, THDCA, GHCA, GHDCA, wMCA, aMCA, bMCA, HCA, and HDCA were categorized as muricholates. Bile acids within each group were summed to calculate the total percentage detected for each group. Two-sample t-tests were used to determine if there were differences in the percentage of these bile acid groups in the total bile acid pool between the serum samples of the Bangladeshi and American cohorts and between serum and stool samples of the Bangladeshi children. EED biomarkers including REG 1B, MPO, CRP, SCD14, LM ratio, and SIBO AUC were separately regressed on categories of bile acids that were identified as significantly different between the American and Bangladeshi populations using univariate linear regression. Finally, we created similar linear regression models to predict anthropometric variables including WAZ, BAZ, LAZ, and WHZ scores.

As an exploratory endeavor, we then conducted a secondary analysis of data that was collected as part of two other cohorts in 2-year-old Bangladeshi children from the same neighborhood in Mirpur, Dhaka. SIBO data, collected using identical methodology to that detailed above, was also available on these samples. 80 paired serum and stool samples collected between May 2011 and March 2016 had bile acid analysis performed by liquid chromatography-mass spectroscopy using the Biocrates AbsoluteIDQ p180 Kit and high-performance liquid chromatography column per manufacturer’s protocols (Biocrates, Inc Aliso Viejo, CA USA). Bile acids were converted into percentages of total bile acids detected. If there were no detectable levels, bile acids were valued at a 0. Then bile acids were grouped based on the bile acid metabolism pathway as follows: CDCA and CA were categorized as unconjugated primary bile acids, TCA, GCA, TCDCA, and GCDCA were categorized as conjugated primary bile acids, UDCA, DCA, and LCA were categorized as unconjugated secondary bile acids, TUDCA, GUDCA, TDCA, GDCA, TLCA, and GLCA were categorized as conjugated secondary bile acids, and TaMCA, TbMCA, aMCA, bMCA, oMCA, and HDCA were categorized as muricholates. Linear regression models were completed between these bile acid categories and SIBO AUC data. Finally, two-sample t-tests were conducted to determine if there were differences between bile acid groups of the 2-year old cohort and the American children, as well as between the 2-year-old cohort and the original Bangladeshi cohort of 6 – 9-month-old children.

The collection and analysis of Bangladeshi samples was approved by the Ethics and Research Review Committees at the icddr,b and the Institutional Review Board at the University of Virginia. Reliance agreements between the University of Virginia and the icddr,b with Virginia Commonwealth University were also established for this study. Informed consent for participation was received by both parents of enrolled children. For the American control group, the study was approved by the Institutional Review Board at Virginia Commonwealth University. A waiver of consent was obtained to use discarded samples.

## Results

The cohort of 100 6–9-month-old Bangladeshi consisted of 51 females and 49 males. The average age was 229 days (range: 185-265 days). The average (and standard deviation) of weight-for-height Z score (WHZ) was −0.75 SD (± 0.69 SD), height-for-age z score (HAZ) was −1.60 SD (± 0.76 SD), weight-for-age z score (WAZ) was −1.55 SD (± 0.47 SD), and BMI-for-age z score (BAZ) was −0.83 SD (± 0.67 SD) (Table 1).

**Table 1.**
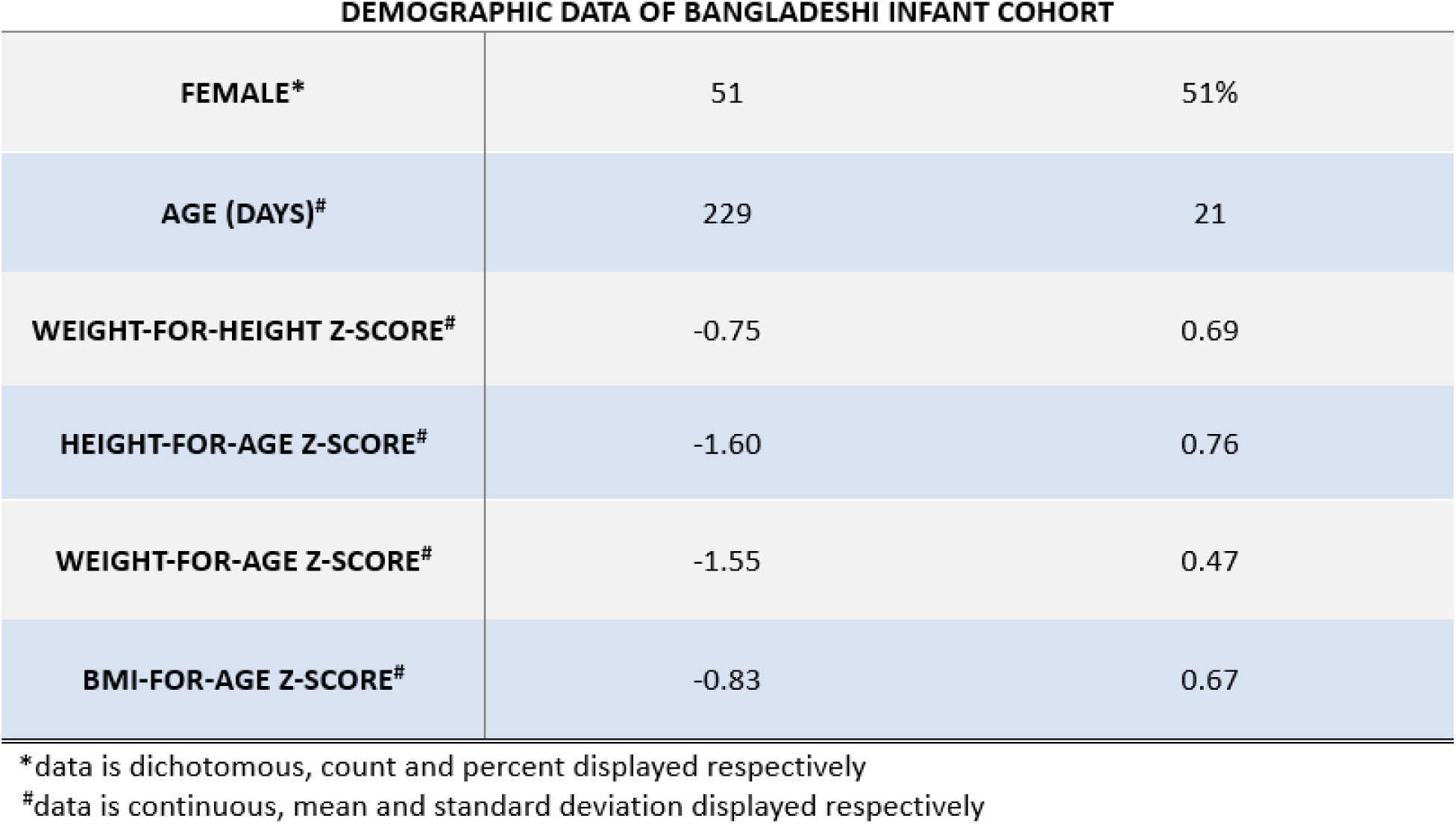
The table presents demographic characteristics of the study population of 100 Bangladeshi infants (age 6-9 months), including sex distribution, age, and anthropometric Z-scores. Sex is provided as both count and percentage. Continuous variables are displayed as means with standard deviations.

| DEMOGRAPHIC DATA OF BANGLADESHI INFANT COHORT |  |  |
| --- | --- | --- |
| **FEMALE\*** | 51 | 51% |
| **AGE (DAYS)#** | 229 | 21 |
| **WEIGHT-FOR-HEIGHT Z-SCORE#** | -0.75 | 0.69 |
| **HEIGHT-FOR-AGE Z-SCORE#** | -1.60 | 0.76 |
| **WEIGHT-FOR-AGE Z-SCORE#** | -1.55 | 0.47 |
| **BMI-FOR-AGE Z-SCORE#** | -0.83 | 0.67 |

\*data is dichotomous, count and percent displayed respectively
#data is continuous, mean and standard deviation displayed respectively

In the serum samples, the Bangladeshi children averaged 216,539.519 nmol/L unconjugated primary bile acids, 1,537.32 nmol/L conjugated primary bile acids, 23,184.71 nmol/L primary sulfated bile acids, 4,412.88 nmol/L secondary bile acids, 93.03 nmol/L secondary conjugated bile acids, 774.41 nmol/L secondary sulfated bile acids, 23,997.97 nmol/L secondary bile acid derivatives, and 14,032.88 nmol/L muricholates. They had a total average bile acid serum sample concentration of 11,956.37 nmol/L. In comparison, the American group has an average serum distribution of 1,096.50 nmol/L unconjugated primary bile acids, 74.03 nmol/L conjugated primary bile acids, 0.00 primary sulfated bile acids, 0.00 secondary bile acids, 56.56 nmol/L secondary conjugated bile acids, 0.00 secondary sulfated bile acids, 20.04 nmol/L secondary bile acid derivatives, and 695.26 nmol/L muricholates. They had a total average bile acid serum sample concentration of 2,569.67 nmol/L. For analysis purposes between the two cohorts, the percentage distribution of each category was calculated and then compared. C4 levels were also obtained in both groups, with Bangladeshi children having an average of 27.72 nmol/L and American children of 0.00 nmol/L (Supplementary Table 1).

As compared to American children, Bangladeshi children’s serum had higher levels of primary unconjugated bile acids (65.23% vs. 44.25%, p = .003), primary sulfated bile acids (12.98% vs. <0.001%, p = .01), and secondary bile acids derivatives (11.35% vs. 0.97%, p = .03). Bangladeshi children had lower percentages of muricholates (6.16% vs. 27.02%, p = <0.001), primary conjugated bile acids (0.69% vs 2.74%, p = <0.001), and secondary conjugated bile acids (0.05% vs. 3.31%, p = <0.001) compared to American children. There was no significant difference in percentage of unconjugated secondary bile acids, percentage of secondary sulfated bile acids, or total bile acids between the two groups (Figure 1). There was no significant difference between percentage of muricholates, unconjugated primary bile acids, conjugated primary bile acids, sulfated primary bile acids, unconjugated secondary bile acids, conjugated secondary bile acids, sulfated secondary bile acids, or secondary bile acid metabolite levels in the stool and serum of the 6 – 9-month-old Bangladeshi children (Figure 2).

**Figure 1.**
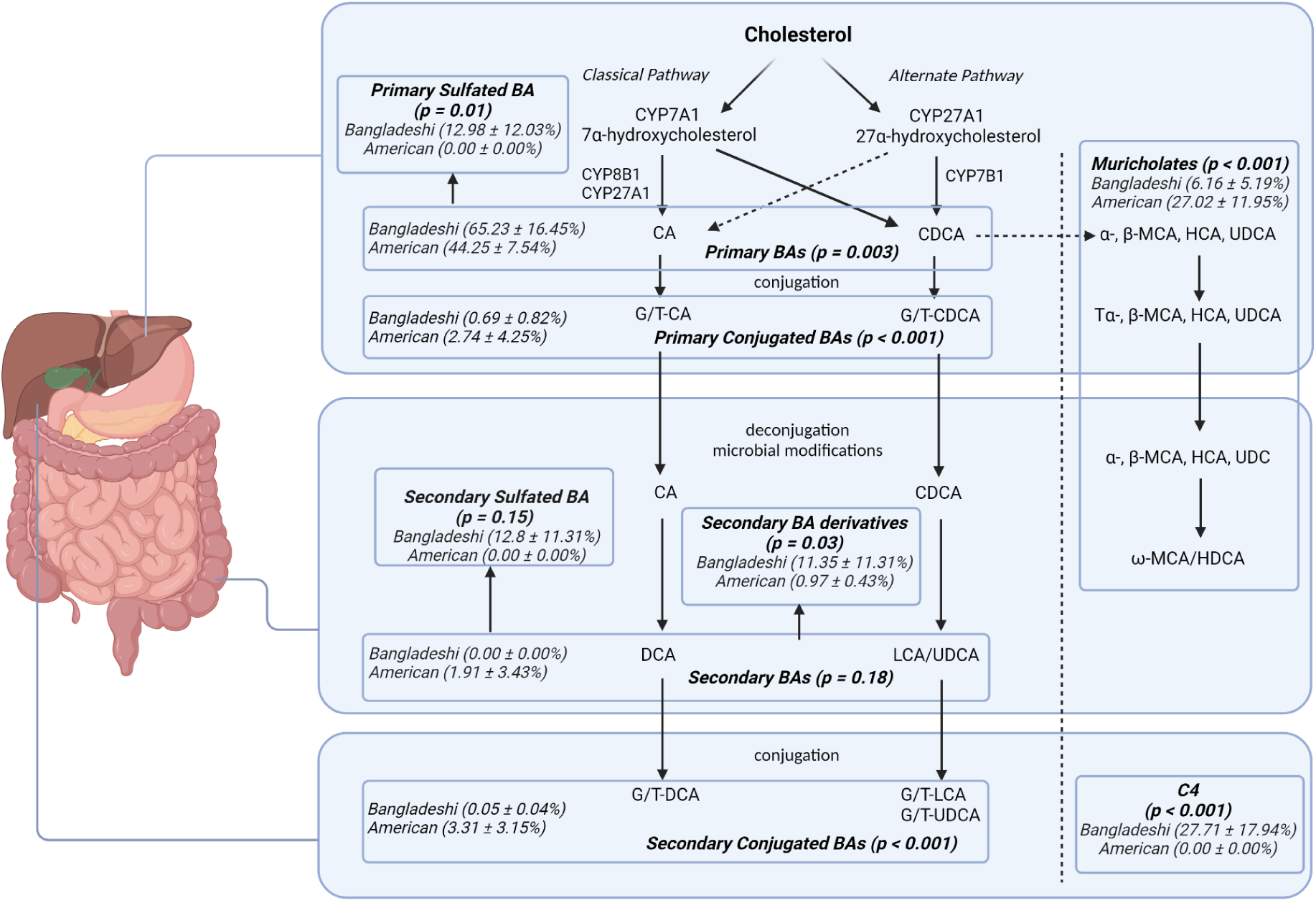
Percent distributions comparing the younger Bangladeshi infants’ serum bile acids and C4 levels to the American children are displayed with mean and standard deviation. Secondary bile acid derivatives include 7-Keto DCA, 7-Keto LCA, 3-Keto LCA, isoDCA, isoLCA, and allo isoLCA. Bangladeshi infants demonstrate a deficiency in primary bile acid conjugation as compared to American children with compensatory increase in hydrophilic primary sulfated bile acids. C4 is also increased.

**Figure 2.**
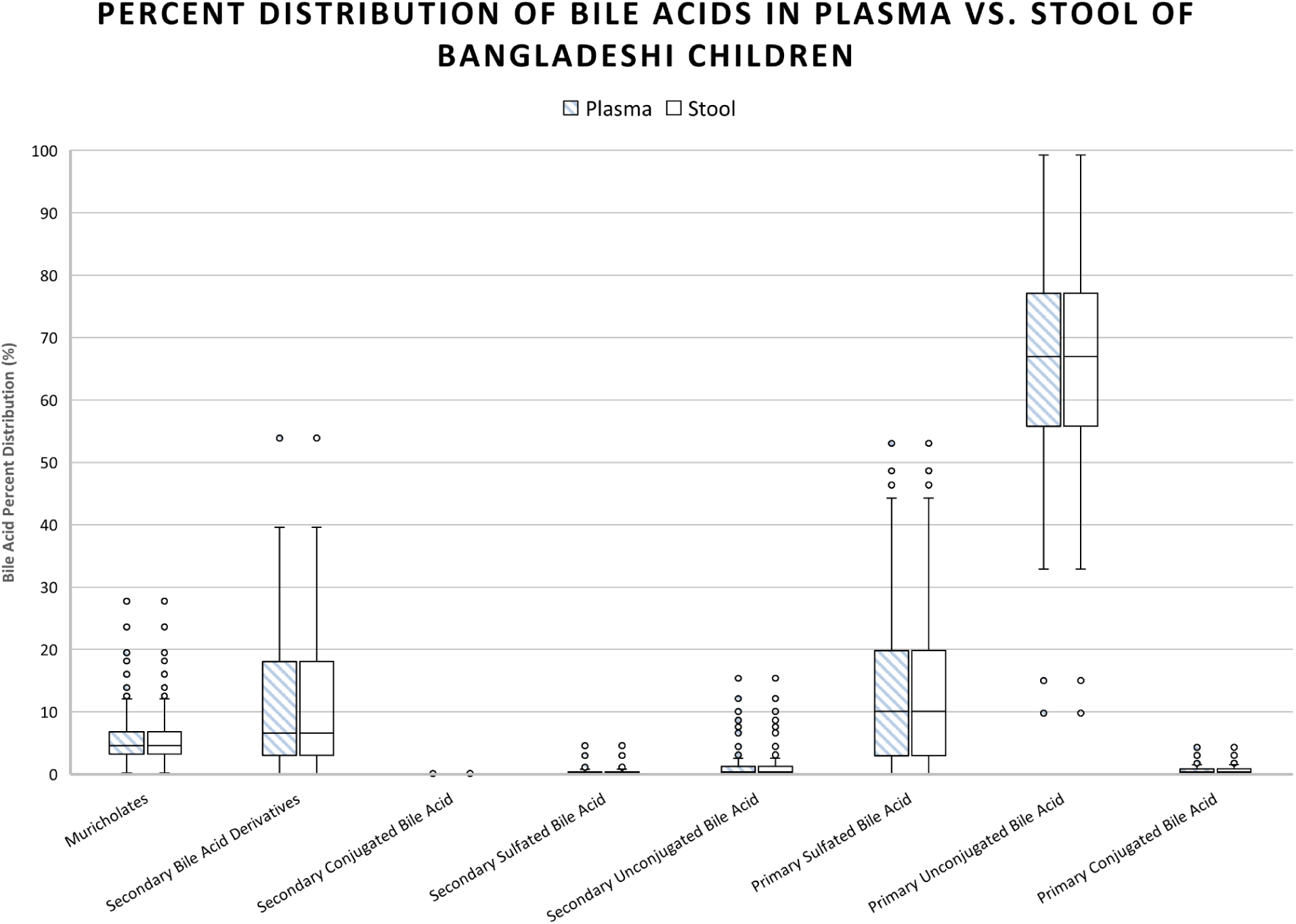
Stool and plasma bile acid pools were compared in Bangladeshi 6 – 9 month olds. Plasma distribution is shown as shaded box-and-whisker plots while stool distribution is illustrated with clear box-and-whisker plots. The central line in each box represents the median, while the box spans the interquartile range (IQR). Whiskers extend to 1.5 times the IQR, with individual points representing outliers. There were no significant differences between the groups in regard to bile acid percent distributions.

In the stool of Bangladeshi children, conjugated secondary bile acids had an inverse relationship with SIBO AUC. (regression coefficient [β] = −1096.68, p = 0.05) while MPO had an inverse association with sulfated secondary bile acids (β = −0.40, p = 0.04) (Figure 3 and 4). No other groups of bile acids were significantly associated with inflammatory biomarkers.

**Figure 3.**
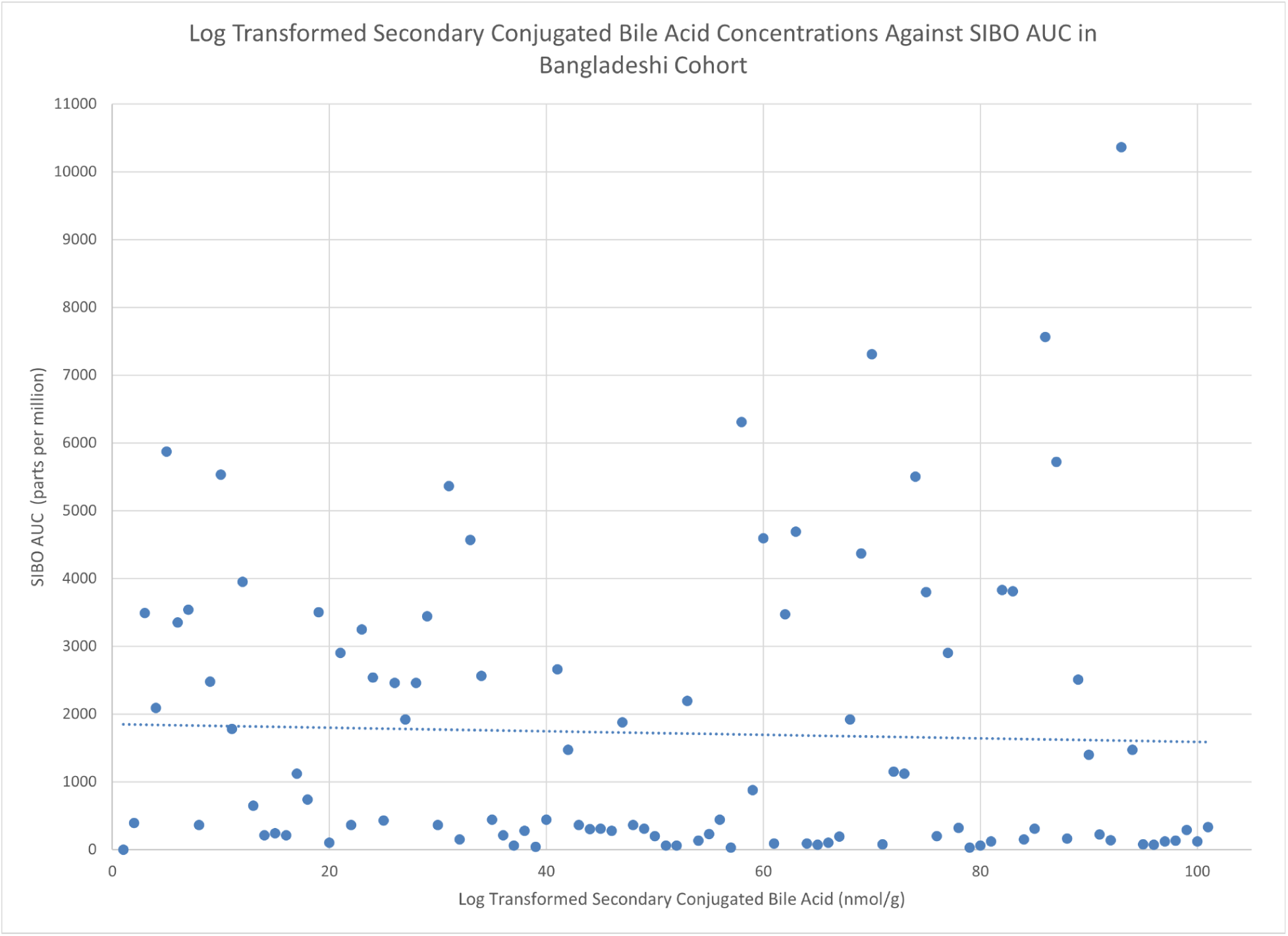
Log transformed secondary conjugated bile acid concentrations were plotted against SIBO AUC in the Bangladeshi infants. SIBO is associated with a decrease in secondary conjugated bile acids (β = −1096.68, p = 0.05).

**Figure 4.**
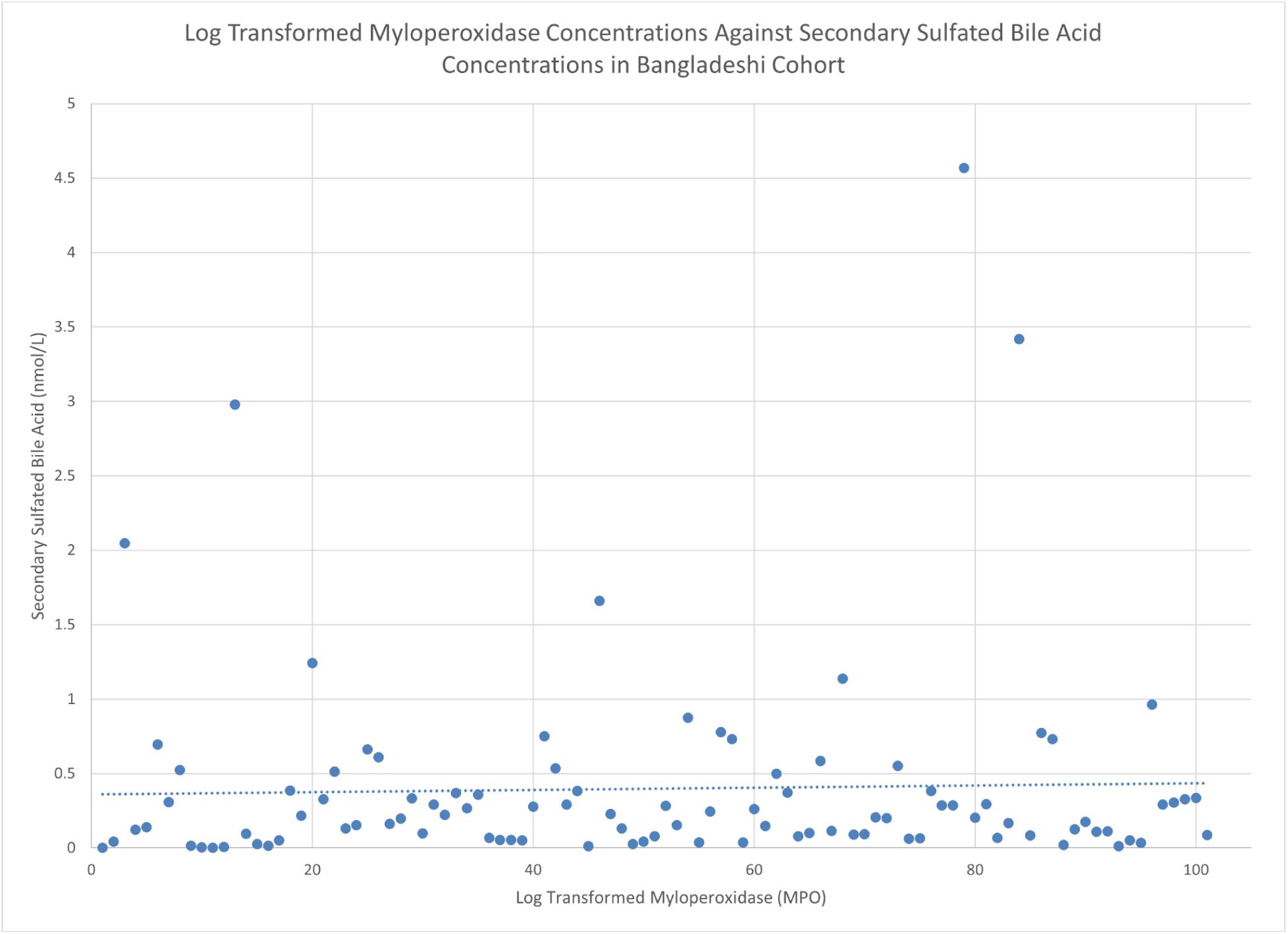
Log transformed myeloperoxidase were regressed on secondary sulfated bile acid concentrations in the Bangladeshi infants. As MPO increases, there is a decrease in the concentration of secondary sulfated bile acids (β = −0.40, p = 0.04).

Unconjugated primary bile acids were associated with weight-for-age (β = −0.27, p = 0.01) and height-for-age z scores (β = −0.01, p = 0.03) (Figure 5, Panel A and B). There was no significant association against weight-for-height or BMI-for-age. Further, C4 concentrations were associated with height-for-age z scores (β = 0.65, p = 0.04) (Figure 5, Panel C). There was no significant association with C4 with weight-for-age, weight-for-height, or BMI-for-age. No other bile acids were significantly associated with weight-for-age, height-for age, weight-for-height, or BMI-for-age.

**Figure 5.**
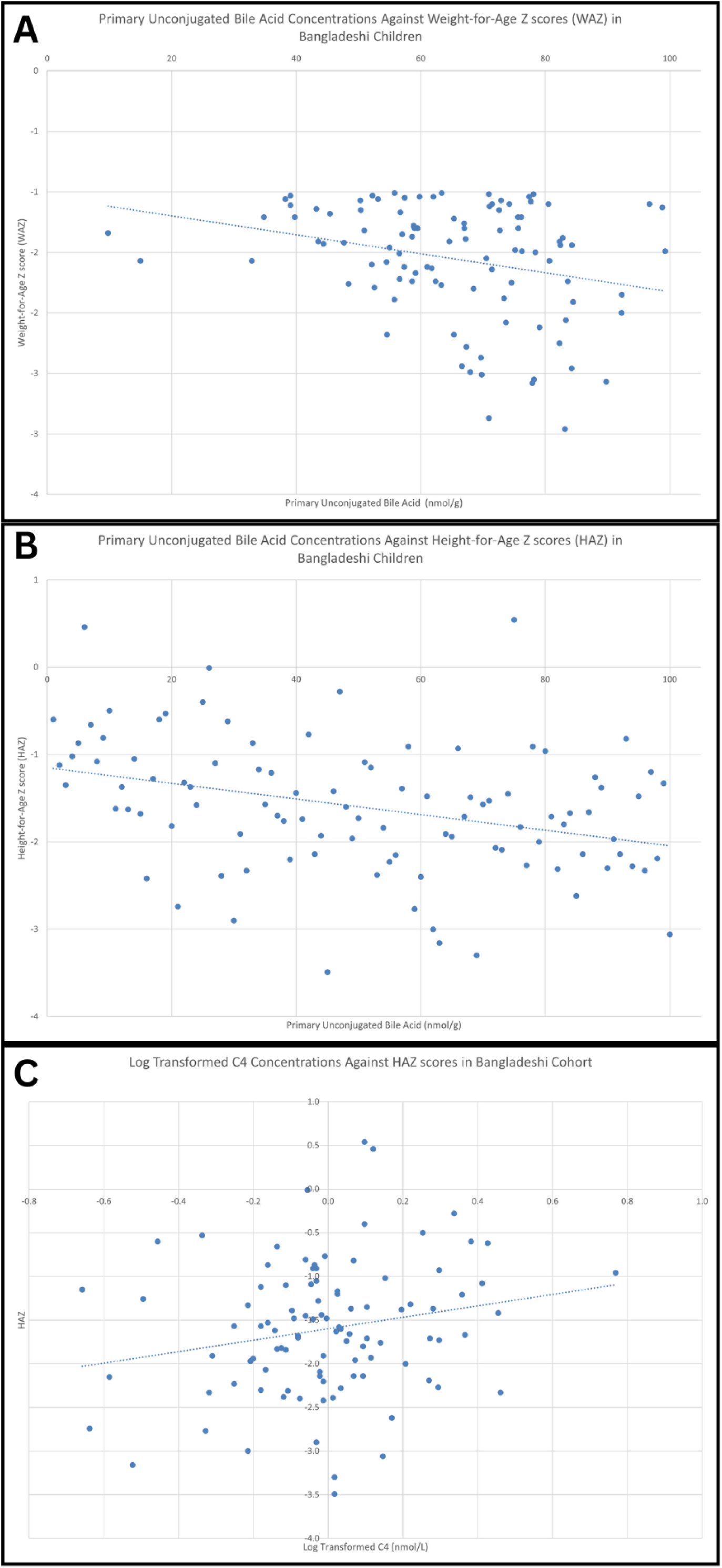
Panel. **A.** Primary unconjugated bile acid concentrations are plotted against WAZ scores in the Bangladeshi infants. There is a significant correlation with drop in WAZ score as primary unconjugated bile acid concentrations increase (β = −0.01, p = 0.01). **Panel B.** Primary unconjugated bile acid concentrations are plotted against HAZ scores in the Bangladeshi infants. There is a significant correlation with drop in HAZ score as primary unconjugated bile acid concentrations increase (β = −0.01, p = 0.03). **Panel C.** Log transformed C4 concentrations were regressed on HAZ scores in the Bangladeshi infants. Regression shows that there is an increase in HAZ with increased C4 levels (β = 0.65, p = 0.04).

The plasma samples of the 6 – 9-month-old Bangladeshi children were compared to the plasma samples of 80 2-year-old Bangladeshi children. As compared to the 2-year-old children, the 6 – 9-month-old Bangladeshi children’s serum had higher levels of unconjugated primary bile acids (65.23% vs. 9.20%, p = <0.001) and muricholates (6.16% vs. 0.26%, p = <0.001). They had lower percentages of primary conjugated bile acids (0.69% vs 80.38%, p = <0.001), and conjugated secondary bile acids (0.05% vs. 7.24%, p = <0.001) compared to the 2-year-old children. There was no significant difference in percentage of unconjugated secondary bile acids between the two groups (Figure 6). There were no significant associations between the older children’s bile acid group percentages and SIBO AUC.

**Figure 6.**
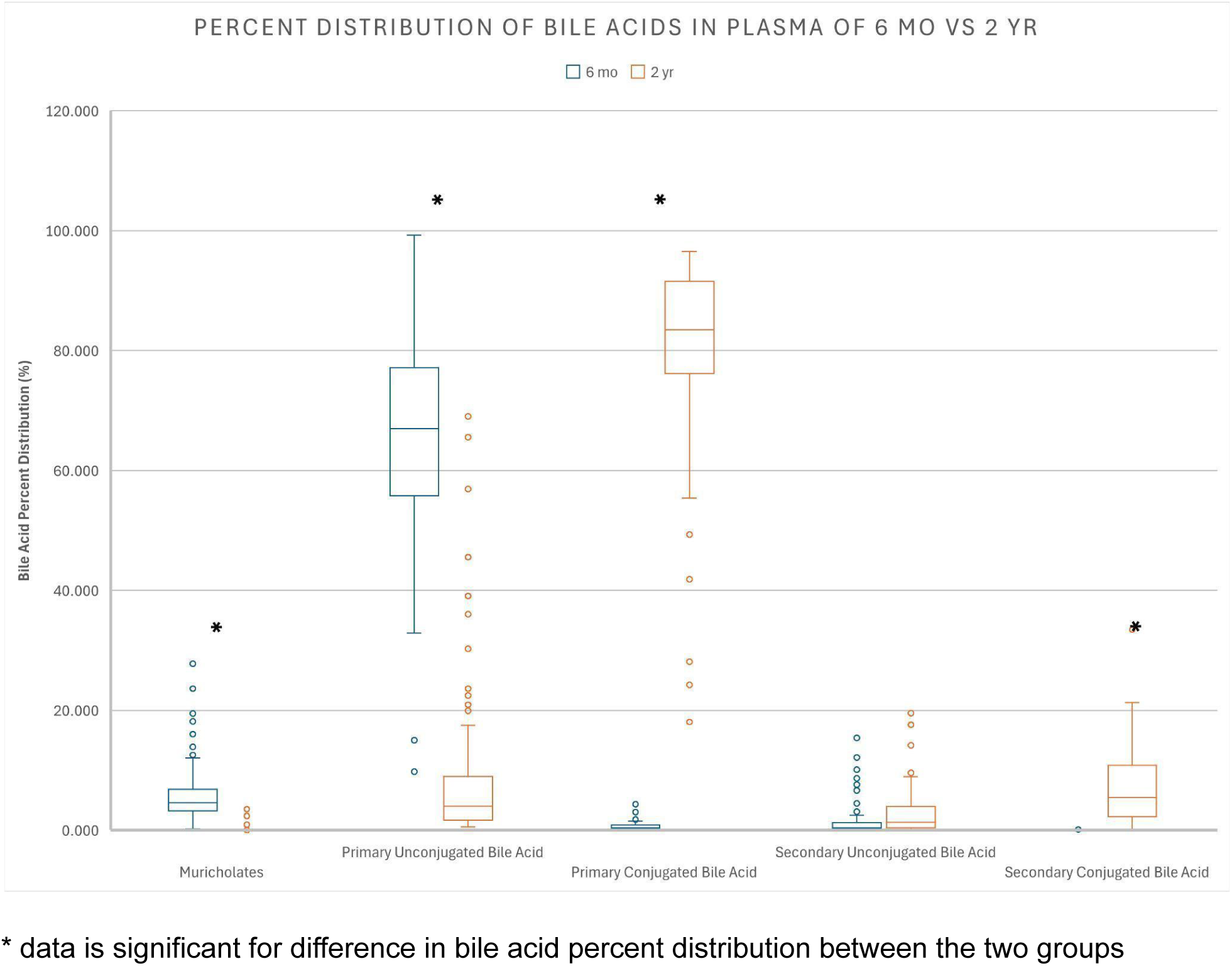
Box-and-whiskers plot showing percent distribution of bile acids in plasma between the Bangladeshi infants (aged 6-9 months) and the Bangladeshi children (aged 2 years old). The younger Bangladeshi infants’ distributions are shown in blue and the older Bangladeshi children are shown in orange. The central line in each box represents the median, while the box spans the interquartile range (IQR). Whiskers extend to 1.5 times the IQR, with individual points representing outliers. The younger infants showed higher levels of unconjugated bile acids compared to their older counterparts.

Finally, when compared to the American children, the 2-year-old Bangladeshi children had higher levels of unconjugated secondary bile acids (2.93% vs. <0.001%, p = <0.001) but lower levels of muricholates (0.26% vs. 27.02%, p = 0.02). There was no significant difference in percentage of unconjugated primary bile acids, conjugated primary bile acids, or conjugated secondary bile acids between the two groups.

## Discussion

Our study demonstrates that dysfunction in bile acid metabolism is associated with stunting and wasting in Bangladeshi children with EED. Overall, when compared to age-matched American children, the Bangladeshi children showed increased levels of unconjugated primary bile acids and sulfated primary bile acids, suggesting arrested metabolism. This dysregulation likely stems from defective hepatic conjugation pathways, as evidenced by significantly reduced levels of primary conjugated bile acids and diminished downstream bile acids. Conjugated bile acids, which are more hydrophilic, play a crucial role in nutrient absorption. Conjugation with glycine or taurine enhances solubility and absorption of hydrophobic nutrients such as long chain fatty acids, cholesterol and fat-soluble vitamins by forming mixed micelles in the small intestine (12). In contrast, hydrophobic bile acids have potent inflammatory properties that can damage the liver, intestine, and other tissues, whereas hydrophilic bile acids exert anti-inflammatory effects (5). Therefore, the observed conjugation defect may contribute to long-term stunting and malnutrition by impairing nutrient absorption and promoting both systemic and enteric inflammation.

Younger Bangladeshi children demonstrated a conjugation defect while older children did not, as shown by the elevated levels of primary and secondary conjugated bile acids in the 2-year-olds. This suggests that impoverished Bangladeshi children have a delay in maturation of their conjugation pathways rather than an intrinsic deficit as seen in patients with bile acid-CoA:amino acid N-acyltransferase (BAAT) and bile acid-CoA ligase (SLC27A5) gene defects (20). This finding corroborates a previous study of Malawian children that inversely correlated levels of CA and CDCA with age in children with EED (19). In a study conducted on microbial challenged mice, protein deficiency led to increased levels of primary bile acids compared to secondary bile acids (3). The dysfunction noted in our cohort of Bangladeshi children may be attributed to their poor nutritional status from an early age, leading to the delay in conjugation that was noted in this study. Another possibility is a microbiota mediated effect. The gut microbiota plays a crucial role in bile acid metabolism, including processes like deconjugation to produce unconjugated bile acids and 7α-dehydroxylation to yield secondary bile acids (16). The gut microbiota also matures over the first 2 years of life with children with EED having a more immature microbiome than expected based on chronological age (21). The gut microbiota’s role in regulating bile acid distribution may explain the differences observed in Bangladeshi children affected by environmental enteric dysfunction (EED).

Younger Bangladeshi children demonstrated very little difference in the bile acid profile patterns between their stool and serum. In normal physiology children reabsorb their bile acids in their ileum for transport back to the liver via portal blood circulation to initiate negative feedback on subsequent bile acid synthesis, and therefore we expected a difference in bile acid levels between stool and serum (16). However, it is possible that the intestinal barrier breakdown associated with EED is sufficient to equilibrate the serum and stool bile acid levels.

Currently, there is no known treatment for EED. Our data suggests correction of the early life conjugation defect could present a therapeutic opportunity. A study targeting defective bile acid amidation due to a deficiency in bile acid CoA:amino acid N-acyl transferase (BAAT) showed improvement in fat-soluble vitamin absorption after treatment with glycocholic acid (GCA) (10). Patients in the study demonstrated a lack of conjugated primary bile acids at baseline, similar to the children in our study. Although enrollment required a genetically confirmed BAAT deficiency, this therapeutic approach may have broader applicability beyond genetically defined cases.

The 100 6 – 9-month-old Bangladeshi children also demonstrated an inverse relationship with increased levels of secondary conjugated bile acids correlating with lower levels of SIBO AUC. Small intestinal bacterial overgrowth is already independently linked to growth stunting (8) but this study explores the potential role of SIBO on bile acid metabolism as a mediating factor. Another study linked patients with malabsorption syndrome with SIBO to significantly elevated levels of unconjugated bile acids compared to those without SIBO (1). This initial correlation warrants further research into how SIBO may affect gut bile acid conjugation and the potential downstream effects on growth and inflammation.

Our study had several notable strengths. First, the study conducted rigorous field collection with concomitant stool and serum samples. Second, we utilized an expanded bile acid profile to understand the accessory pathways being utilized in Bangladeshi children in the absence of effective primary bile acid conjugation. The inclusion of a wide range of bile acids in the analysis, including sulfated forms and muricholates, provides a more detailed understanding of bile acid metabolism and their implications. Finally, we sampled American children of similar age as a high-income country control.

This study also had several notable limitations that should be considered when interpreting its findings. First, as a cross-sectional study, it lacks longitudinal data on the enrolled children. Although we included a separate cohort of older children from the same impoverished neighborhood, these participants were not the same individuals as those in the younger cohort. This discrepancy limits our ability to assess individual-level influences or developmental trajectories in bile acid metabolism over time. Another key limitation is the relatively small sample size of the American cohort. While the comparison provides valuable insights, the limited size and diversity of the American group may not fully capture the heterogeneity present in a broader population. This highlights a broader gap in the literature, as few studies have attempted to characterize normal variations in bile acid metabolism across genetically and socioeconomically diverse pediatric populations at different developmental stages.

This study identified a correlation between conjugation defects and growth deficits in young Bangladeshi children. Further research is needed to better understand the impact of bile acid metabolic dysfunction in impoverished children from low-income countries and to explore potential avenues for novel interventions aimed at mitigating growth impairments.

## Funding

Funding for this work was provided by grants from the Children’s Hospital Foundation at VCU and National Institutes of Health (1K23HD097282 to JD, 5R01AI043596 to WAP), and the Bill and Melinda Gates Foundation (OPP1017093 to WP). This study was partially supported by VA Merit Award 5I01BX005730, VA ShEEP grants (1 IS1 BX004777-01), National Institutes of Health Grant 2R56DK115377-05A1, PIDS Summer Research Scholars Award, and VCU SOM Dean’s Summer Research Fellowship. Dr. Zhou is the recipient of a Research Career Scientist Award from the Department of Veterans Affairs (IK6BX004477).

## Data Availability

All data produced in the present study are available upon reasonable request to the authors

## Acknowledgements

Authors would like to acknowledge the children and families of Mirpur Dhaka without whom this research would not have been possible. icddr,b acknowledges its core donors, the Government of Bangladesh, and the Government of Canada for providing unrestricted support and commitment to icddr,b’s research effort.

---

**Supplementary Table 1.** The table presents bile acid distributions in serum samples of the Bangladeshi infants and the American cohort. Distribution is given as mean and standard deviation.

| **BILE ACID DISTRIBUTIONS IN SERUM SAMPLES OF BANGLADESHI\* AND AMERICAN COHORTS** |  |  |  |  |
| --- | --- | --- | --- | --- |
|  | Bangladeshi |  | American |  |
| **PRIMARY UNCONJUGATED BILE ACIDS** | 216,539.51 | 546,833.52 | 1,096.50 | 624.28 |
| **PRIMARY CONJUGATED BILE ACIDS** | 1,537.32 | 2,270.37 | 74.03 | 120.60 |
| **PRIMARY SULFATED BILE ACIDS** | 23,184.71 | 22,600.43 | 0.00 | 0.00 |
| **SECONDARY UNCONJUGATED BILE ACIDS** | 44,12.88 | 9,275.94 | 0.00 | 0.00 |
| **SECONDARY CONJUGATED BILE ACIDS** | 93.03 | 147.59 | 56.56 | 34.00 |
| **SECONDARY SULFATED BILE ACIDS** | 774.41 | 1,472.08 | 0.00 | 0.00 |
| **SECONDARY BILE ACID METABOLITES** | 23,997.97 | 30,034.10 | 20.04 | 4.11 |
| **MURICHOLATES** | 14,032.88 | 17,747.84 | 695.26 | 476.95 |
| **TOTAL BILE ACIDS** | 11,956.37 | 22,801.49 | 2,569.67 | 1,520.02 |
| **C4** | 27.71 | 17.94 | 0.00 | 0.00 |

\* Bangladeshi cohort of 100 infants aged 6-9 months
Note: data displayed as mean and standard deviation, all units in nmol/L

## Notes

### Competing Interest Statement

The authors have declared no competing interest.

## References

1. Bala L, Ghoshal UC, Ghoshal U, et al. Malabsorption syndrome with and without small intestinal bacterial overgrowth: A study on upper-gut aspirate using 1H NMR spectroscopy. Magnetic Resonance in Medicine. 2006;56(4):738–744. doi:10.1002/mrm.21041

2. Bartelt LA, Bolick DT, Guerrant RL. Disentangling Microbial Mediators of Malnutrition: Modeling Environmental Enteric Dysfunction. Cell Mol Gastroenterol Hepatol. 2019;7(3):692–707. doi:10.1016/j.jcmgh.2018.12.006

3. Bhatt AP, Arnold JW, Awoniyi M, et al. Giardia Antagonizes Beneficial Functions of Indigenous and Therapeutic Intestinal Bacteria during Malnutrition. bioRxiv. Published online January 23, 2024:2024.01.22.575921. doi:10.1101/2024.01.22.575921

4. Chen RY, Kung VL, Das S, et al. Linking the duodenal microbiota to stunting in a cohort of undernourished Bangladeshi children with enteropathy. N Engl J Med. 2020;383(4):321–333. doi:10.1056/NEJMoa1916004

5. Chiang JYL. Bile Acid Metabolism and Signaling. Compr Physiol. 2013;3(3):1191–1212. doi:10.1002/cphy.c120023

6. Church JA, Rukobo S, Govha M, et al. Associations between biomarkers of environmental enteric dysfunction and oral rotavirus vaccine immunogenicity in rural Zimbabwean infants. eClinicalMedicine. 2021;41. doi:10.1016/j.eclinm.2021.101173

7. Donowitz JR, Haque R, Kirkpatrick BD, et al. Small Intestine Bacterial Overgrowth and Environmental Enteropathy in Bangladeshi Children. mBio. 2016;7(1):e02102–02115. doi:10.1128/mBio.02102-15

8. Donowitz JR, Pu Z, Lin Y, et al. Small Intestine Bacterial Overgrowth in Bangladeshi Infants Is Associated With Growth Stunting in a Longitudinal Cohort. Am J Gastroenterol. 2022;117(1):167–175. doi:10.14309/ajg.0000000000001535

9. Guerrant RL, DeBoer MD, Moore SR, Scharf RJ, Lima AAM. The impoverished gut—a triple burden of diarrhoea, stunting and chronic disease. Nat Rev Gastroenterol Hepatol. 2013;10(4):220–229. doi:10.1038/nrgastro.2012.239

10. Heubi JE, Setchell KDR, Jha P, et al. Treatment of Bile Acid Amidation Defects with Glycocholic Acid. Hepatology. 2015;61(1):268–274. doi:10.1002/hep.27401

11. Korpe PS, Petri WA. Environmental Enteropathy: Critical implications of a poorly understood condition. Trends Mol Med. 2012;18(6):328–336. doi:10.1016/j.molmed.2012.04.007

12. Marschall HU, Beuers U. When Bile Acids Don’t Get Amidated. Gastroenterology. 2013;144(5):870–873. doi:10.1053/j.gastro.2013.03.018

13. Mehta HC, Saini AS, Singh H, Dhatt PS. Biochemical aspects of malabsorption in marasmus. Br J Nutr. 1984;51(1):1–6. doi:10.1079/bjn19840003

14. Moreau GB, Ramakrishnan G, Cook HL, et al. Childhood growth and neurocognition are associated with distinct sets of metabolites. EBioMedicine. 2019;44:597–606. doi:10.1016/j.ebiom.2019.05.043

15. Musa MA, Kabir M, Hossain MI, et al. Measurement of intestinal permeability using lactulose and mannitol with conventional five hours and shortened two hours urine collection by two different methods: HPAE-PAD and LC-MSMS. PLoS One. 2019;14(8):e0220397. doi:10.1371/journal.pone.0220397

16. Ramírez-Pérez O, Cruz-Ramón V, Chinchilla-López P, Méndez-Sánchez N. The Role of the Gut Microbiota in Bile Acid Metabolism. Annals of Hepatology. 2017;16:S21–S26. doi:10.5604/01.3001.0010.5672

17. Rosenberg IH, Hardison WG, Bull DM. Abnormal Bile-Salt Patterns and Intestinal Bacterial Overgrowth Associated with Malabsorption. New England Journal of Medicine. 1967;276(25):1391–1397. doi:10.1056/NEJM196706222762501

18. Schneider RE, Viteri FE. Luminal events of lipid absorption in protein-calorie malnourished children; relationship with nutritional recovery and diarrhea. I. Capacity of the duodenal content to achieve micellar solubilization of lipids. The American Journal of Clinical Nutrition. 1974;27(8):777–787. doi:10.1093/ajcn/27.8.777

19. Semba RD, Gonzalez-Freire M, Moaddel R, et al. Environmental Enteric Dysfunction Is Associated With Altered Bile Acid Metabolism. J Pediatr Gastroenterol Nutr. 2017;64(4):536–540. doi:10.1097/MPG.0000000000001313

20. Setchell KDR, Heubi JE, Shah S, et al. Genetic Defects in Bile Acid Conjugation Cause Fat-Soluble Vitamin Deficiency. Gastroenterology. 2013;144(5):945–955.e6. doi:10.1053/j.gastro.2013.02.004

21. Subramanian S, Huq S, Yatsunenko T, et al. Persistent gut microbiota immaturity in malnourished Bangladeshi children. Nature. 2014;510(7505):417–421. doi:10.1038/nature13421

22. Tickell KD, Atlas HE, Walson JL. Environmental enteric dysfunction: a review of potential mechanisms, consequences and management strategies. BMC Medicine. 2019;17(1):181. doi:10.1186/s12916-019-1417-3

23. Vonaesch P, Araújo JR, Gody JC, et al. Stunted children display ectopic small intestinal colonization by oral bacteria, which cause lipid malabsorption in experimental models. Proc Natl Acad Sci U S A. 2022;119(41):e2209589119. doi:10.1073/pnas.2209589119

24. Wang Y, Tai YL, Zhao D, et al. Berberine Prevents Disease Progression of Nonalcoholic Steatohepatitis through Modulating Multiple Pathways. Cells. 2021;10(2):210. doi:10.3390/cells10020210

25. Zhang L, Voskuijl W, Mouzaki M, et al. Impaired Bile Acid Homeostasis in Children with Severe Acute Malnutrition. PLoS One. 2016;11(5):e0155143. doi:10.1371/journal.pone.0155143

26. Zhao X, Setchell KDR, Huang R, et al. Bile Acid Profiling Reveals Distinct Signatures in Undernourished Children with Environmental Enteric Dysfunction. J Nutr. 2021;151(12):3689–3700. doi:10.1093/jn/nxab321

